# Statin Therapy in Ischemic Stroke Patients with Atrial Fibrillation: Efficacy and Safety Outcomes

**DOI:** 10.1101/2024.04.19.24306111

**Authors:** RAF and RAF NOAC-Investigators, Michele Marvardi, Mario Ferrante, Maurizio Paciaroni, Azmil Abdul-Rahim, Georgios Tsivgoulis, David Julian Seiffge, Stefan T. Engelter, Philippe Lyrer, Alexandros A. Polymeris, Tolga Dittrich, Virginia Cancelloni, Annaelle Zietz, Gian Marco De Marchis, Jukka Putaala, Daniel Strbian, Liisa Tomppo, Patrik Michel, Davide Strambo, Alexander Salerno, Peter Vanacker, Susanna Zuurbier, Laetitia Yperzeele, Caroline M.J. Loos, Manuel Cappellari, Andrea Emiliani, Marialuisa Zedde, Cecilia Becattini, Rosario Pascarella, Jesse Dawson, Robert Cronshaw, Erika Schirinzi, Massimo Del Sette, Christoph Stretz, Narendra Kala, Ashley Schomer, Brian Mac Grory, Mahesh Jayaraman, Shadi Yaghi, Karen L. Furie, Luca Masotti, Elisa Grifoni, Danilo Toni, Angela Risitano, Anne Falcou, Luca Petraglia, Enrico Maria Lotti, Lucia Pavolucci, Piergiorgio Lochner, Giorgio Silvestrelli, Alfonso Ciccone, Andrea Alberti, Michele Venti, Ilaria Leone De Magistris, Maria Giulia Mosconi, Michela Giustozzi, Odysseas Kargiotis, Alessandro Rocco, Marina Diomedi, Simona Marcheselli, Kateryna Antonenko, Eugenia Rota, Tiziana Tassinari, Valentina Saia, Francesco Palmerini, Paolo Aridon, Valentina Arnao, Serena Monaco, Salvatore Cottone, Antonio Baldi, Cataldo D’Amore, Walter Ageno, Samuela Pegoraro, George Ntaios, Anastasia Adamou, Dimitrios Sagris, Sotirios Giannopoulos, Maria Kosmidou, Evangelos Ntais, Michele Romoli, Leonardo Pantoni, Silvia Rosa, Pierluigi Bertora, Alberto Chiti, Isabella Canavero, Carlo Emanuele Saggese, Maurizio Plocco, Elisa Giorli, Lina Palaiodimou, Eleni Bakola, Fabio Bandini, Antonio Gasparro, Valeria Terruso, Marina Mannino, Alessandro Pezzini, Raffaele Ornello, Simona Sacco, Nemanja Popovic, Umberto Scoditti, Antonio Genovese, Yuriy Flomin, Michelangelo Mancuso, Roberto D’Agliano, Marco Baldini, Leonardo Ulivi, Nicola Giannini, Kostantinos Vadikolias, Chrysoula Liantinioti, Maria Chondrogianni, Panagiotis Halvatsiotis, Efstathia Karagkiozi, George Athanasakis, Kostantinos Makaritsis, Alessia Lanari, Turgut Tatlisumak, Monica Acciarresi, Gianni Lorenzini, Rossana Tassi, Francesca Guideri, Maurizio Acampa, Giuseppe Martini, Sung-Il Sohn, Nicola Mumoli, Davide Carrara, Miriam Maccarrone, Franco Galati, Vanessa Gourbali, Giovanni Orlandi, Martina Giuntini, Francesco Corea, Marta Bellesini, Theodore Karapanayiotides, Laszló Csiba, Lilla Szabó, Davide Imberti, Alessio Pieroni, Kristian Barlinn, Lars-Peder Pallesen, Jessica Barlinn, Boris Doronin, Vera Volodina, Giancarlo Agnelli, Dirk Deleu, Bruno Bonetti, Luana Gentile, Giuseppe Reale, Pietro Caliandro, Andrea Morotti, Vieri Vannucchi, Marina Padroni, Federica Letteri, Mauro Magoni, Cindy Tiseo, Alberto Rigatelli, Aurelia Zauli, Francesco Bossi, Valeria Caso

**Author notes:** **Corresponding author:** Michele Marvardi, MD Stroke Unit and Division of Cardiovascular Medicine, University of Perugia, Italy 06121, Perugia, Italy.

## Abstract

**Background:** The efficacy and safety of statins for secondary prevention in patients who have experienced a cardioembolic stroke are not well-defined. However, previous observational data reported hyperlipidemia as a risk factor for both ischemic and bleeding complications in patients with AF and previous stroke. Based on these premises, we conducted a sub-analysis of the RAF and RAF-NOAC studies to evaluate the efficacy and safety of statins in secondary prevention in patients with acute ischemic stroke and AF.

**Methods:** We combined patient data from the RAF and RAF-NOAC studies, prospective observational studies conducted across Stroke Units in Europe, the United States, and Asia from January 2012 to June 2016. We included consecutive patients with AF who suffered an acute ischemic stroke with a follow-up of 90 days. Our outcomes were the combined endpoint, including stroke, transient ischemic attack, systemic embolism, symptomatic intracerebral hemorrhage, and major extracranial bleeding. Furthermore, both ischemic and hemorrhagic outcomes were evaluated separately.

**Results:** A total of 1.742 patients were included (46% male), and 898 (52%) received statins after the index event. At multivariable analysis, statin use was statistically associated with age (OR 0.92, 95% CI 0.97 - 0.99, p = 0.001), male sex (OR 1.35, 95% CI 1.07 - 1.70, p = 0.013), anticoagulation (OR 2.53, 95% CI 1.90 - 3.36, p <0.0001), hyperlipidemia (OR 5.52, 95% CI 4.28 - 7.12, p <0.0001), paroxysmal AF (OR 1.40, 95% CI 1.12 - 1.75, p = 0.003), leukoaraiosis (OR 1.39, 95% CI 1.11 - 1.75, p = 0.004) and heart failure (OR 0.72, 95% CI 0.53 - 0.98, p = 0.034). Statin use was not associated with the combined outcome event (OR 0,81, 95% CI 0,56 – 1,19, p = 0.286) and ischemic outcome event (OR 1,13, 95% CI 0,71 – 1,82, p = 0.604) while was associated with a lower risk of hemorrhagic outcome event (OR 0,49, 95% CI 0,27 – 0,88, p = 0.016).

**Conclusions:** Our data show that statins seem to protect against global bleeding events in cardioembolic stroke patients; this may be due to the pleiotropic effect of statins. More data are warranted to confirm these findings.

## Background

While randomized controlled trials (RCTs) have evaluated statins for their potential in preventing recurrent acute ischemic stroke related to atherosclerosis, including large vessel occlusion (LVO) and small vessel disease (SVD), their effectiveness for ischemic stroke of cardioembolic origin remains unclear^1-4^ Although there is a well-documented link between high LDL cholesterol and increased risk of acute ischemic stroke due to LVO and SVD, the association with cardioembolic stroke is not yet clear.^3-7 **8-10**^ The RENo and RENo-ICH studies identified hyperlipidemia as a statistically significant risk factor for both ischemic and hemorrhagic recurrences in patients with cardioembolic stroke. ^11,12^ Additionally, the RENo study revealed that approximately one-third of recurrent events were caused by LVO and SVD rather than cardioembolism. ^11^ Notably, statins have demonstrated efficacy in reducing the risk of such non-cardioembolic events. ^11 13,14^

Beyond their role in lowering cholesterol, statins are known for their benefits in plaque stabilization, endothelial protection, and anti-inflammatory and anti-thrombotic effects—which may theoretically reduce the risk of acute ischemic stroke from any causes. ^15 16^

Despite the benefits, there is a described risk of increased bleeding events, particularly intracerebral, reported mainly in secondary prophylaxis. Two large RCTs including patients with non-cardioembolic stroke, the Heart Protection Study (HPS) and the Stroke Prevention by Aggressive Reduction in Cholesterol Levels (SPARCL) Study, showed a reduction in the number of cerebral ischemic events in the group of statin-treated patients but a statistically significant increase of bleeding events was observed, also in secondary prophylaxis. ^8,9^

Based on these premises, we conducted a sub-analysis of the RAF and RAF-NOAC studies to evaluate the efficacy and safety of statins in secondary prevention in patients with ischemic stroke and AF.

## Methods

The data that supported the findings of this study are available from the corresponding author upon reasonable request.

We merged the datasets from the RAF and RAF-NOAC studies, both prospective observational studies. ^17,18^ The RAF study was conducted from January 2012 to March 2014 in 29 Stroke Units, while the RAF-NOAC study spanned from April 2014 to June 2016 in 35 Stroke Units throughout Europe, the United States, and Asia. ^17,18^ Both studies recruited patients who experienced an acute ischemic stroke and were diagnosed with AF, either prior to or following their stroke, with no contraindications to anticoagulant therapy. Ethical approval for both studies was obtained from local Institutional Review Boards as necessary.

The stroke severity was evaluated upon admission using the NIH Stroke Scale (NIHSS). All patients underwent either a non-contrast CT or an MR scan to rule out intracranial hemorrhage. Revascularization treatments, following the local standard protocols, were administered when indicated. Standard care in the Stroke Unit, along with monitoring and treatment, adhered to the international guidelines for acute ischemic stroke. Decisions regarding the selection and initiation timing of anticoagulants for secondary prevention were at the discretion of the attending physicians. The RAF study encompassed patients prescribed vitamin K antagonists or NOACs, whereas the RAF-NOAC study was exclusive to those treated with NOACs. Comprehensive medical history, clinical assessment, admission ECG, and 48-hour ECG monitoring post-index stroke were conducted to identify non-valvular AF, which was then classified accordingly:

- Paroxysmal: associated with episodes terminating spontaneously within seven days;
- Persistent: associated with episodes lasting more than seven days or requiring pharmacological and/ or electrical cardioversion;
- Permanent: persisting for more than one year, either because cardioversion had failed or had not been pursued.

### Risk factors

We collected data on established stroke risk factors, including age, gender, and medical histories of hypertension, diabetes mellitus, smoking, hyperlipidemia, ischemic heart disease, peripheral arterial disease, alcohol abuse, obesity, and prior stroke or transient ischemic attack (TIA). Hypertension was defined by a blood pressure reading of ≥140/90 mmHg on at least two occasions prior to the stroke or current antihypertensive medication use. Diabetes mellitus was identified by fasting glucose levels

≥126 mg/dL, postprandial levels ≥200 mg/dL, HbA1c ≥6.5%, or ongoing antidiabetic treatment. Hyperlipidemia at admission was defined by total cholesterol ≥200 mg/dL, triglycerides ≥140 mg/dL, or existing lipid-lowering therapy. Symptomatic ischemic heart and peripheral arterial diseases were confirmed through clinical history and diagnostic procedures.

Additionally, we evaluated white matter changes, categorizing leukoaraiosis (defined on the first computed tomography examination as ill-defined and moderately hypodense areas of ≥5 mm) on initial CT or MR imaging as absent or present, regardless of severity. ^19^

Baseline measurements at admission for all patients included fasting serum glucose, cholesterol profiles (total, HDL, and LDL), platelet count, coagulation times, and blood pressure.

Physicians prescribed statins based on clinical judgment from among the seven commercially available in Europe, the United States, and Asia. We also documented the use of antiplatelet, anticoagulant, and thrombolytic agents before and during the study.

### Outcomes

Primary study outcome at 90 days was defined as the composite of, ischemic stroke, transient ischemic attack, symptomatic systemic embolism, symptomatic intracerebral hemorrhage, and major extracranial hemorrhage. Additionally, ischemic and hemorrhagic events were analyzed separately. Stroke was defined as the sudden onset of a new focal neurological deficit of a vascular origin in a site consistent with the territory of a major cerebral artery, and categorized as either ischemic or hemorrhagic. Hemorrhagic transformations revealed on neuroimaging 24-72h after onset were not considered outcome events, unless they were classified as being symptomatic. TIA was defined as a transient episode of neurological dysfunction caused by focal brain ischemia without acute infarction. Systemic embolism was defined as an acute vascular occlusion of an extremity or organ confirmed at imaging, surgery, or autopsy. Major extracerebral bleeding was defined as a reduction in the hemoglobin level of at least 2g per deciliter, requiring a blood transfusion of at least 2U, or symptomatic bleeding in a critical area or organ.

### Statistical analyses

For patients with or without statins after the index event and with or without primary outcome event at 90 days, differences in clinical characteristics and risk factors were evaluated using the χ2 test of proportions (with a 2-sided α level of 5%), by calculating odds ratio (OR) and 95% confidence intervals (CIs). Multivariable logistic regression analysis was performed to identify independent associations with statin use and outcome events. The variables included in this analysis were the following: age, sex, oral anticoagulant therapy, diabetes, hypertension, hyperlipidemia, paroxysmal atrial fibrillation, history of stroke/TIA, alcoholism, heart failure, history of myocardial infarction, peripheral arterial disease, leukoaraiosis, and smoking (CHA2DS2-VASc and total cholesterol were excluded due to the concurrent presence within the multivariate analysis of the individual factors already included in the CHA2DS2-VASc score and hyperlipidemia). Subsequent analyses were conducted to identify independent predictive factors for both the Ischemic Outcome and the Hemorrhagic Outcome events, including the same predictive factors used previously.

Finally, we carried out Cox Regression analyses (Cox proportional hazard model) to study in detail the effect of statins over the 90 days of follow-up on the Hemorrhagic Outcome and symptomatic intracranial hemorrhage; the variables included in these two analyses were the same: statin therapy, hyperlipidemia, paroxysmal atrial fibrillation, and CHA2DS2-VASc ≥ 4. The results of the Cox Regression are reported in Hazard Ratios (HR).

## Results

In total, the study enrolled 1,742 patients. After excluding 34 due to incomplete follow-up data, the analysis included 1,708 participants. Among them, 811 (46.6%) were male and with a mean age of 76+/-9.86.

In this cohort of 1,708 patients, 898 (52%) were prescribed statins after the index event. Atorvastatin was the most common, used by 28.5% of patients, followed by Simvastatin at 12.8% and Rosuvastatin at 5.6%. A smaller number (0.6%) were on Pravastatin. Fluvastatin and Pitavastatin were the least used, with only one patient on each. Data on statin usage was missing for 69 (4%). Only 13 (0.7%) of patients received Ezetimibe, excluded in the statistical analyses.

Regarding anticoagulant therapy, 1,380 (79.2%) patients received treatment, 841 (48.3%) on NOACs, and 539 (30.9%) on Warfarin. The remaining 361 (20.7%) were not on oral anticoagulants, most commonly used subcutaneous low molecular weight heparin or, less frequently, antiplatelet drugs alone. In Table 1 are listed the main risk factors.

**Table 1:** Clinical characteristics of the patients enrolled in the study.

|  | Patients (n:1742) |
| --- | --- |
| **Age (y)** | 76 ± 9,86 |
| **Sex (male)** | 811 (46,6%) |
| **Oral anticoagulants (OAC)** | 1380 (79,2%) |
| **NOAC** | 841 (48,3%) |
| **Warfarin** | 539 (30,9%) |
| **No OAC** | 361 (20,7%) |
| **Diabetes** | 396 (22,7%) |
| **Hypertension** | 1343 (77,1%) |
| Hyperlipidemia | 575 (33%) |
| AF classification |  |
| paroxysmal | 732 (42%) |
| persistent | 307 (17,6%) |
| permanent | 692 (39,7%) |
| History of stroke/TIA | 456 (26,2%) |
| Alcoholism | 121 (6,9%) |
| Heart failure | 311 (17,8%) |
| History of myocardial infarction | 255 (14,6%) |
| Peripheral arterial disease | 149 (8,5%) |
| Leukoaraiosis | 861 (49,4%) |
| CHA<sub>2</sub>DS<sub>2</sub>-VASc ≥ 4 Before or after??? | 1580 (90,6%) |
| Tobacco use (current or previous) | 535 (30,7%) |
| Total cholesterol (mg/dL) | 180,9 ± 44,3 |
| Statins | 898 (52%) |
| Atorvastatin | 497 (28,5%) |
| Rosuvastatin | 97 (5,6%) |
| Simvastatin | 223 (12,8%) |
| Pravastatin | 10 (0,6%) |
| Fluvastatin | 1 (0,1%) |
| Pitavastatin | 1 (0,1%) |
| Unspecified statin | 69 (4%) |

**Table 2:** Multivariate Analysis: Predictors for Statin Therapy.

|  | OR | 95% IC | P |
| --- | --- | --- | --- |
| Age (y) | 0,98 | 0,97 – 0,99 | **0,001** |
| Sex (male) | 1,35 | 1,07 -1,70 | **0,013** |
| Oral anticoagulants (OAC) | 2,53 | 1,90 – 3,36 | **<0,0001** |
| Diabetes | 1,24 | 0,95 – 1,61 | 0,116 |
| Hypertension | 1,23 | 0,94 – 1,60 | 0,130 |
| Hyperlipidemia | 5,52 | 4,28 – 7,12 | **<0,0001** |
| Paroxysmal AF | 1,40 | 1,12 – 1,75 | **0,003** |
| History of stroke/TIA | 1,08 | 0,84 – 1,39 | 0,569 |
| Alcoholism | 1,44 | 0,93 – 2,25 | 0,106 |
| Heart failure | 0,72 | 0,53 – 0,98 | **0,034** |
| History of myocardial infarction | 1,27 | 0,91 – 1,78 | 0,158 |
| Peripheral arterial disease | 0,71 | 0,47 – 1,08 | 0,109 |
| Leukoaraiosis | 1,39 | 1,11 – 1,75 | **0,004** |
| Tobacco use (current or previous) | 0,87 | 0,67 – 1,12 | 0,281 |

**Table 3:**
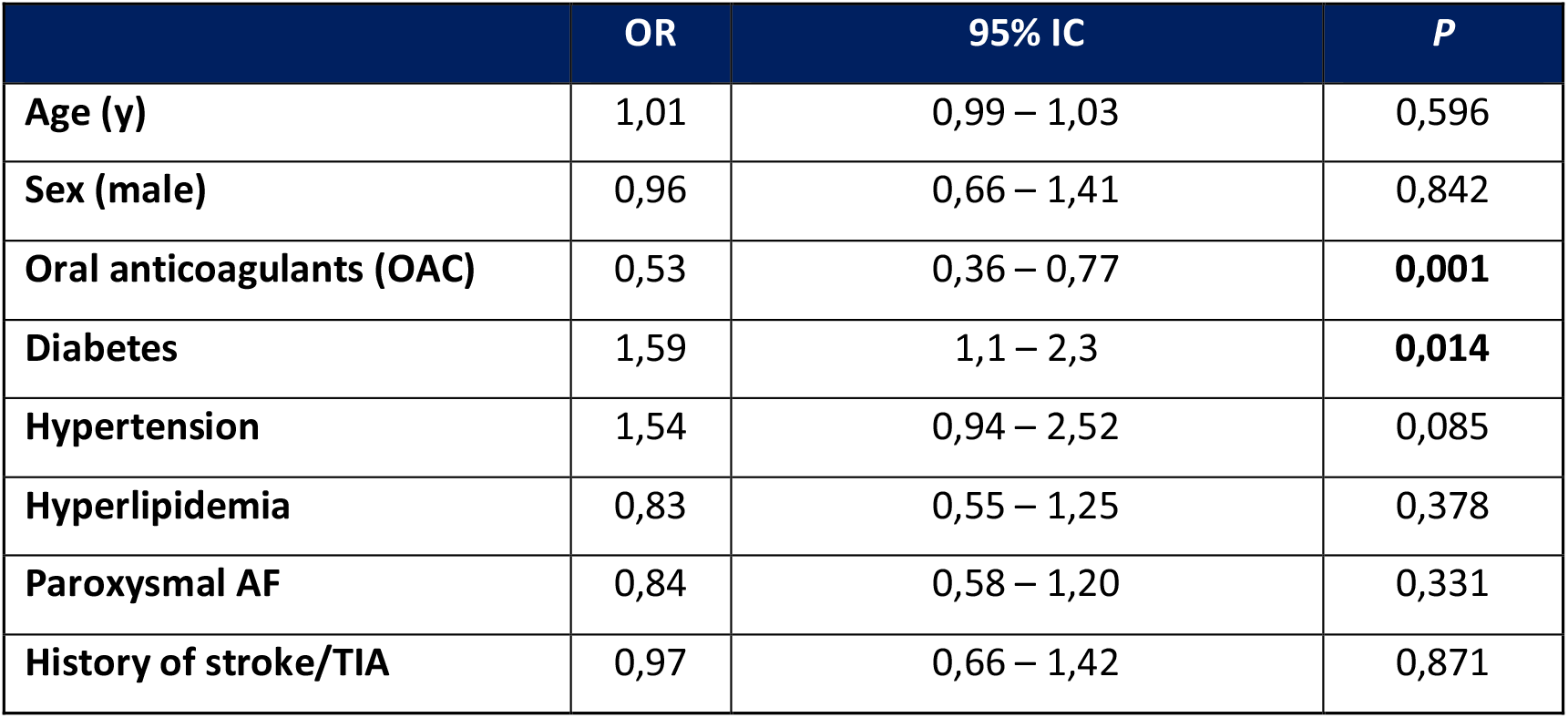
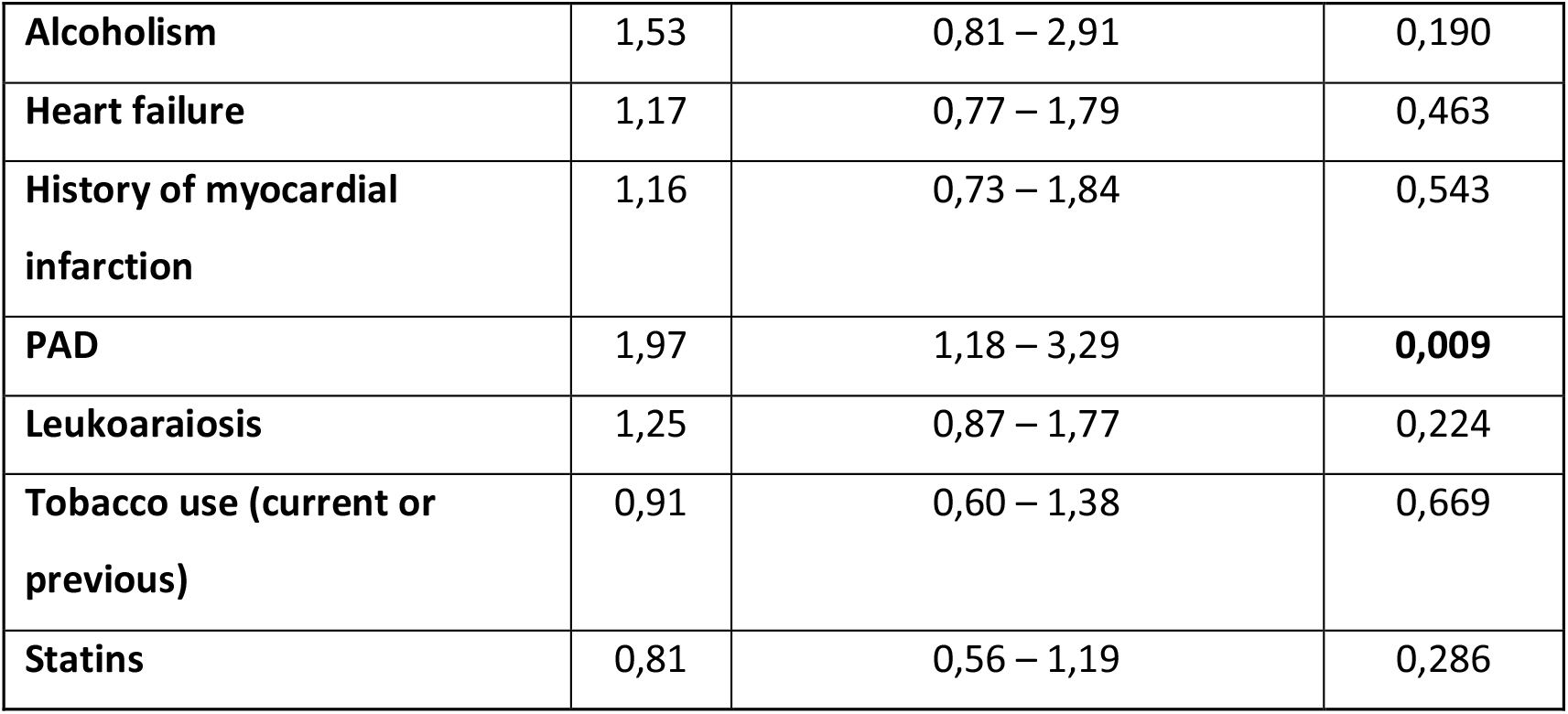
Multivariate Analysis: Predictors for Combined Outcome.

|  | OR | 95% IC | P |
| --- | --- | --- | --- |
| Age (y) | 1,01 | 0,99 – 1,03 | 0,596 |
| Sex (male) | 0,96 | 0,66 – 1,41 | 0,842 |
| Oral anticoagulants (OAC) | 0,53 | 0,36 – 0,77 | **0,001** |
| Diabetes | 1,59 | 1,1 – 2,3 | **0,014** |
| Hypertension | 1,54 | 0,94 – 2,52 | 0,085 |
| Hyperlipidemia | 0,83 | 0,55 – 1,25 | 0,378 |
| Paroxysmal AF | 0,84 | 0,58 – 1,20 | 0,331 |
| History of stroke/TIA | 0,97 | 0,66 – 1,42 | 0,871 |
| Alcoholism | 1,53 | 0,81 – 2,91 | 0,190 |
| Heart failure | 1,17 | 0,77 – 1,79 | 0,463 |
| History of myocardial infarction | 1,16 | 0,73 – 1,84 | 0,543 |
| PAD | 1,97 | 1,18 – 3,29 | **0,009** |
| Leukoaraiosis | 1,25 | 0,87 – 1,77 | 0,224 |
| Tobacco use (current or previous) | 0,91 | 0,60 – 1,38 | 0,669 |
| Statins | 0,81 | 0,56 – 1,19 | 0,286 |

**Table 4:**
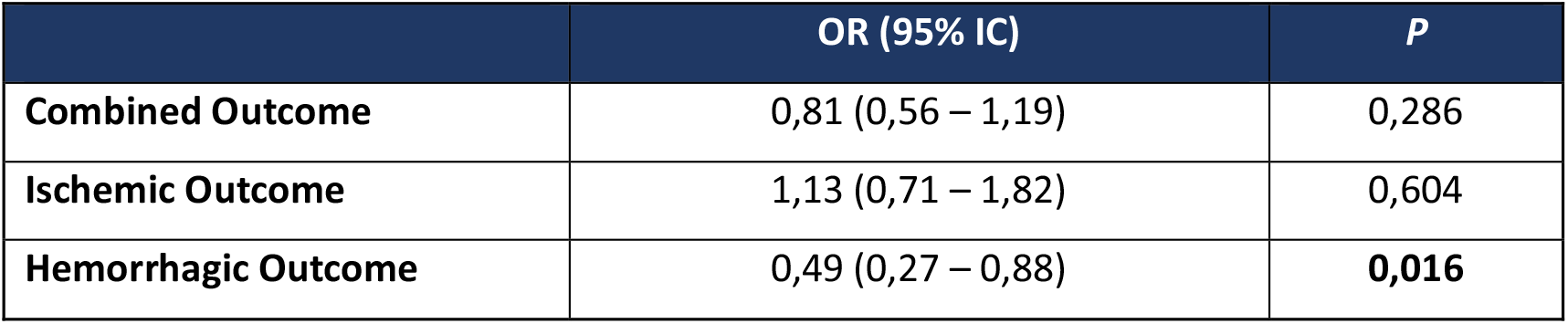
Multivariate analysis on the role of statins on the combined, ischemic and hemorrhagic outcome.

### Predictors for statin therapy

The clinical characteristics of the patients treated and not treated with statins are listed in Table S1. Statin therapy was more frequent in younger patients (74.28±9.07 vs 77.20±10.49, p <0.0001), those taking oral anticoagulants (57% vs 39%, p <0.0001), in patients with hyperlipidemia (50.0% vs 15.4%, p <0.0001), in those with paroxysmal AF (46.8% vs 37.0%, p <0.0001), and in those with a history of myocardial infarction (18.2% vs 10.9%, p <0.0001). Statistical significance was also shown in diabetic patients (25.7% vs 19.5%, p = 0.002), hypertensive patients (80.2% vs 72.8%, p = 0.003), in those with a history of cerebral ischemic events (acute ischemic stroke/TIA, 28.6% vs 23.6%, p = 0.022), in those with leukoaraiosis on imaging studies (52.1% vs 46.6%, p = 0.024), and in smokers, current or past (32.7% vs 28.6%, p = 0.037).

The multivariable analysis identified oral anticoagulation (OR 2.53, 95% CI 1.90 - 3.36, p <0.0001) and hyperlipidemia (OR 5.52, 95% CI 4.28 - 7.12, p <0.0001) as predictors of statin therapy. Conversely, age had an inverse association with statin use (OR 0.98, 95% CI 0.97 - 0.99, p = 0.001), as did heart failure (OR 0.72, 95% CI 0.53 - 0.98, p = 0.034). In male patients, there was an increase in the likelihood of statin therapy (OR 1.35, 95% CI 1.07 - 1.70, p = 0.013). Statins were also more commonly prescribed among patients with paroxysmal atrial fibrillation (OR 1.40, 95% CI 1.12 - 1.75, p = 0.003) and leukoaraiosis (OR 1.39, 95% CI 1.11 - 1.75, p = 0.004).

### Predictors for primary and secondary outcomes

A total of 134 patients (7.8%) experienced a primary outcome, whereas ischemic outcome was observed in 99 individuals (5.8%) and hemorrhagic outcomes in 70 individuals (4.1%). Symptomatic intracerebral hemorrhage alone was reported in 49 patients (2.9%).

A significant positive correlation among primary outcome and age was present: 78.0±8.92 versus 75.71±9.92 (p = 0.005). Also, diabetes was associated with primary outcome: 33.5% versus 21.6% (p = 0.001). Similarly, hypertension (86.0% vs 76.3%, p = 0.006) and PAD (15.2% vs 7.7%, p = 0.003) were statistically significant for primary outcome. Conversely, OAC was inversely associated with the primary outcome: 30.5% versus 49.2% (p = 0.029). Statin therapy also suggested a protective trend (44.5% vs 51.9%), but this did not reach statistical significance (p = 0.084) (Table S2).

On multivariable analysis, OAC was inversely correlated with primary outcome (OR 0.53, 95% CI 0.36 - 0.77, p = 0.001). Conversely, diabetes (OR 1.59, 95% CI 1.1 - 2.3, p = 0.014) and PAD (OR 1.97, 95% CI 1.18 - 3.29, p = 0.009) were associated with an increased risk of primary outcome. Statin use was associated with a reduced risk of primary outcome, but this association did not reach statistical significance (OR 0.81, 95% CI 0.53 - 1.19, p: 0.286).

Regarding secondary outcomes, the hemorrhagic event rate was lower in the statin group (2.9% compared to 5.4%, p = 0.010), as was the rate of symptomatic intracerebral hemorrhage (2.1% vs 3.7%, p=0.043).

On multivariable analysis, statin use was associated with a 51% reduction of total bleeding events, within 90 days (OR 0.49, 95% CI 0.27 - 0.88, p = 0.016) and this effect remains stable over the 90 days as shown in the Cox survival curve analysis (Figure 1). Regarding the symptomatic intracerebral hemorrhage outcome, statin therapy showed an early protective effect with a stable trend over the 90 days of follow-up, albeit in the absence of adequate statistical significance (p = 0.066) (Figure 2).

**Figure 1.**
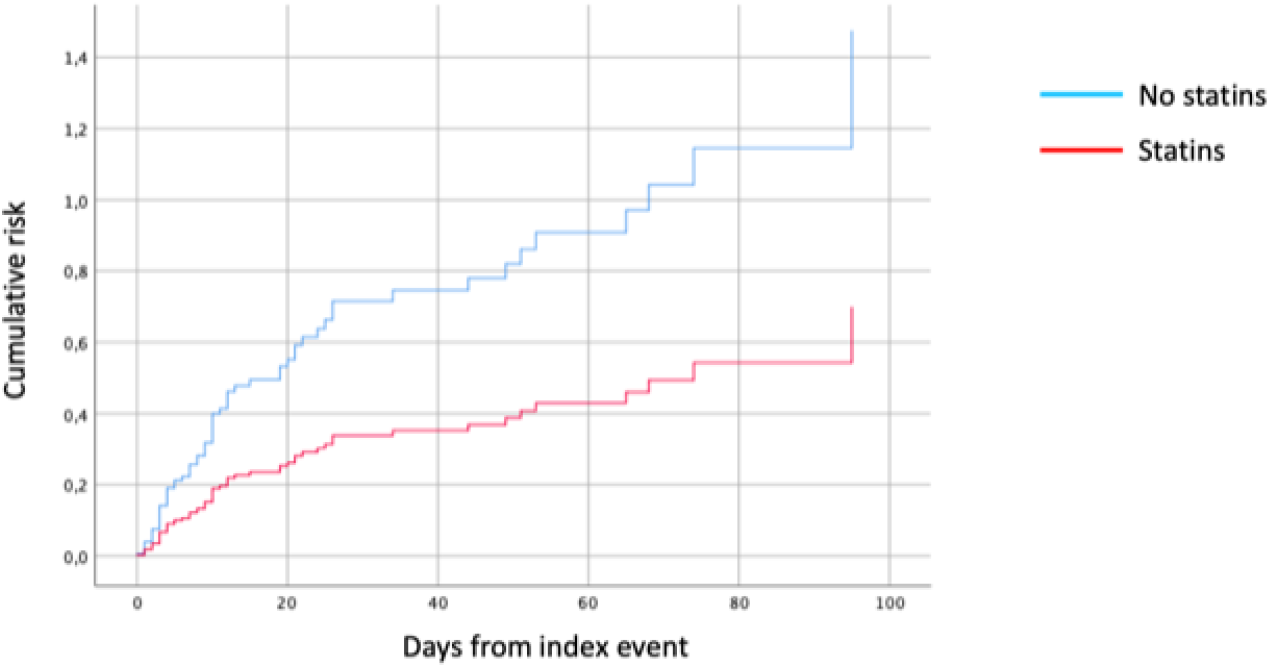
Cox Regression: Statins as an Independent Predictive Factor for Hemorrhagic Outcome.

**Figure 2.**
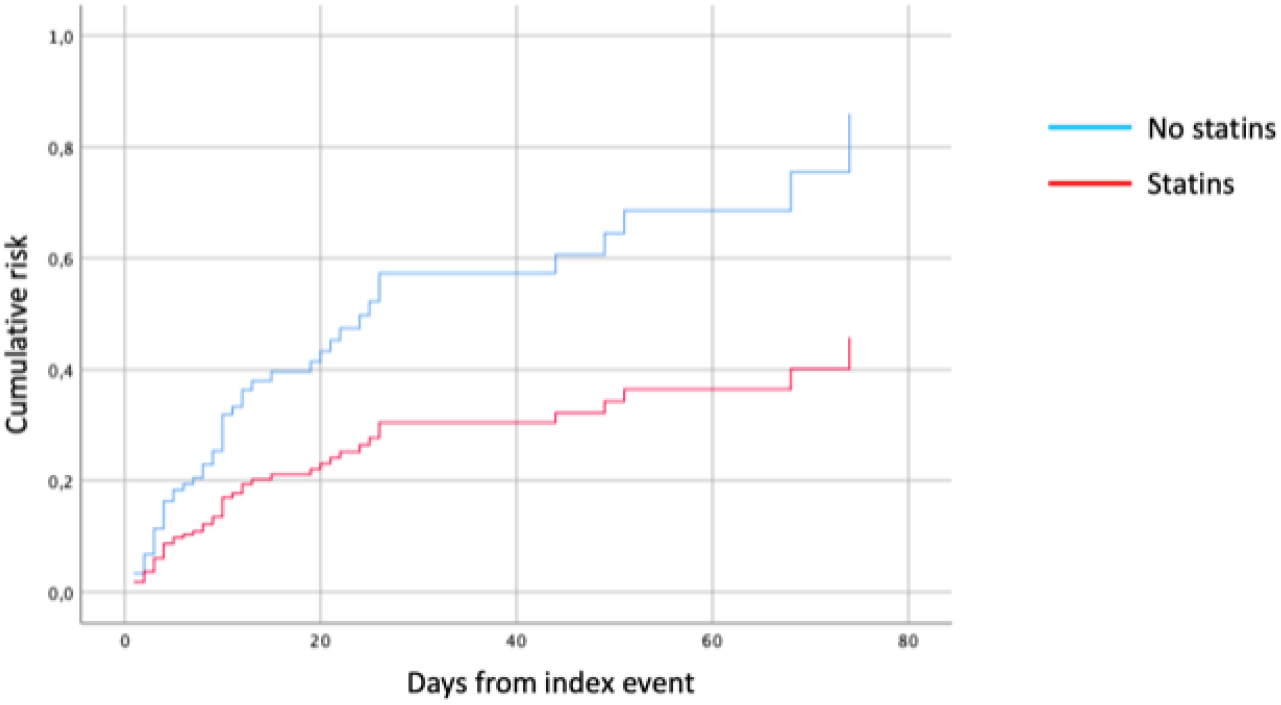
Cox Regression: Statins as an Independent Predictor for Symptomatic Intracranial Hemorrhage.

## Discussion

In this observational study, which included a cohort of patients with AF in secondary prevention for ischemic stroke, statin therapy showed a statistically significant protective effect against global bleeding events during the 90 days following the index cerebral ischemic event. This correlation is probably due to a protective effect of statins against intracerebral hemorrhage. Whereas, for the Combined Outcome, statin therapy showed a protective trend, albeit without adequate statistical significance.

Our findings diverge from those of the Heart Protection Study and the SPARCL Trial, which both observed a statistically significant increase in bleeding events, particularly intracerebral hemorrhage.^8 9^ A subsequent meta-analysis which included data from 24 RCTs, including HPS and the SPARCL Trial, supported a trend towards increased bleeding events in patients undergoing secondary prophylaxis, with RR of 1.73 (95% CI 1.19 – 2.50). ^6^ This analysis suggested that reduced LDL cholesterol could contribute to the increased risk of bleeding. However, it is important to note that data from the HPS and SPARCL trials predominantly influenced this result. ^8 9^ Specifically, in the HPS, the incidence of ICH was 32 cases (21 in the Simvastatin group versus 11 in the placebo group). Given the small sample size, this did not confer statistical significance (RR 1.91, 95% CI 0.92 – 3.95), suggesting that the association may be less robust than initially perceived.^8^ In a sub-analysis of the SPARCL trial, which pooled patients who had an ICH, it was observed that ICH risk was higher in those having a hemorrhagic stroke as the entry event (HR 5.65, 95% CI 2.82 - 11.30, p <0.001) without statistical interactions between statin therapy. ^20^ Consequently, the patients identified as having the highest risk for developing intracranial bleeding were those with a history of a previous bleeding event.

Data from comprehensive national registries in Denmark, ^21-23^ South Korea ^24^and China, ^25^ encompassing nearly 1.5 million patients, consistently reported the safety of statins in preventing cerebral hemorrhagic events, both as primary and secondary prophylaxis. ^21 22,23,25^ Furthermore, the findings suggest that both the duration and the dosage of statin therapy contribute to a protective effect against intracerebral hemorrhage. Patients with prolonged statin use and those on higher dosages had a lower risk of ICH, challenging the previously hypothesized biological mechanism linking LDL reduction to an increased bleeding risk. ^26^ These results are in line with those of an extensive meta-analysis published in 2012 for a total of 31 RCTs with 182,803 patients.^27^ A similar meta-analysis by Hackman et al., which included published and unpublished data from 23 randomized trials and 19 observational studies, reported that statins were not associated with an increased risk of ICH neither in randomized trials (RR 1.10, 95% CI 0.86-1.41) or cohort studies (RR 0.94, 95% CI 0.81-1.10), both in primary (48% of patients included in RCTs) and in secondary (52% of patients included in RCTs) stroke prevention.^28 29^

A recent Japanese study examining a patient cohort similar to ours investigated the impact of statins on individuals with AF who were also on oral anticoagulants. The Authors observed that the group treated with statins, compared to those not on statins, had a higher presence of paroxysmal AF, hypertension, diabetes, and dyslipidemia.^30^ Notably, the statin group demonstrated a reduced risk of major bleeding. Furthermore, this group also showed decreased risks in all-cause mortality, ischemic events, and various types of strokes.^30^

It is plausible that statins protect cerebral arterial vessels (particularly small vessels) from subacute damage due to hypertension, diabetes, and other harmful agents (such as reactive oxygen species, proinflammatory cytokines, etc.) due to their systemic anti-inflammatory and endothelium-protective effects. ^15^

1. The reduction of cellular infiltration and metalloproteinase synthesis and the increase in collagen synthesis guarantee greater stability of the atheromatous plaque preventing plaque ulceration or, if it were already present, prevents its rupture with consequent acute thrombosis and vessel occlusion.
2. Anti-inflammatory effects are mainly due to the reduction of the synthesis of pro-inflammatory cytokines, in particular the C-Reactive Protein, and the NF-kB complex. The systemic anti-inflammatory effect consequently reduces the oxidation of LDL, which underlies atherogenesis.
3. Statins also have an important protective effect on the vascular endothelium: they stimulate the production and release of nitric oxide by endothelial cells, which stimulates vasodilation and protects against endothelial damage thanks to its antioxidant properties.
4. Finally, statins enhance fibrinolysis by stimulating the production and release of t-PA, inhibit platelet aggregation by blocking the release of thromboxane in the blood stream and inhibit the extrinsic pathway of coagulation by reducing the expression of the tissue factor; this effect is added to that of any other antithrombotic ongoing therapies, and prevents thrombosis on plaque with occlusion of the vessel lumen which is the basis of major cardiovascular events.

The cumulative effects of statins may serve as a preventive measure against the onset or progression of cerebral small vessel disease. By safeguarding the integrity of small cerebral vessels, statins potentially mitigate the risk of ischemic events caused by thrombotic occlusions and hemorrhagic events due to the rupture of compromised vessels. Building on the same biological hypothesis, it is plausible that administering statins during the acute phase of an ischemic stroke could lower the likelihood of hemorrhagic transformation.

We did not perform logistic regression for the symptomatic intracranial hemorrhage outcome, due to the small sample size.

This study has several limitations:

- it is an observational study, based on data collected from databases of previous studies, in which neither the use nor the dosage of statins were randomized;
- the duration and intensity of the therapy, the type of statin and the LDL cholesterol levels at entry (index cerebral ischemic event) and at the time of the study outcome events were not considered;
- except for anticoagulants, additional medications that patients were taking besides statins were not included. As a result, we have no information on drug interactions that can potentially enhance or decrease the effects of statins;
- the 90-day follow-up is probably too limited to make conclusions, further studies are needed, both to confirm the safety of statin therapy in the acute and subacute phase, and subsequently to verify whether safety is maintained over a medium-long period of time.

However, this study has a large sample size which gives significant statistical value to the results of the analyses. It also has a prospective design and reflects the real-life experiences of many centers located on different continents (Europe, North America and Asia).

## Conclusions

In this observational study, including patients with AF in secondary prevention for ischemic stroke, statin therapy showed a statistically significant protective effect against global bleeding events (intracranial and extracranial) during the 90 days following the index cerebral ischemic event. This correlation is mainly due to the protective effect of statins on an intracranial hemorrhage.

---

**Table S1:**
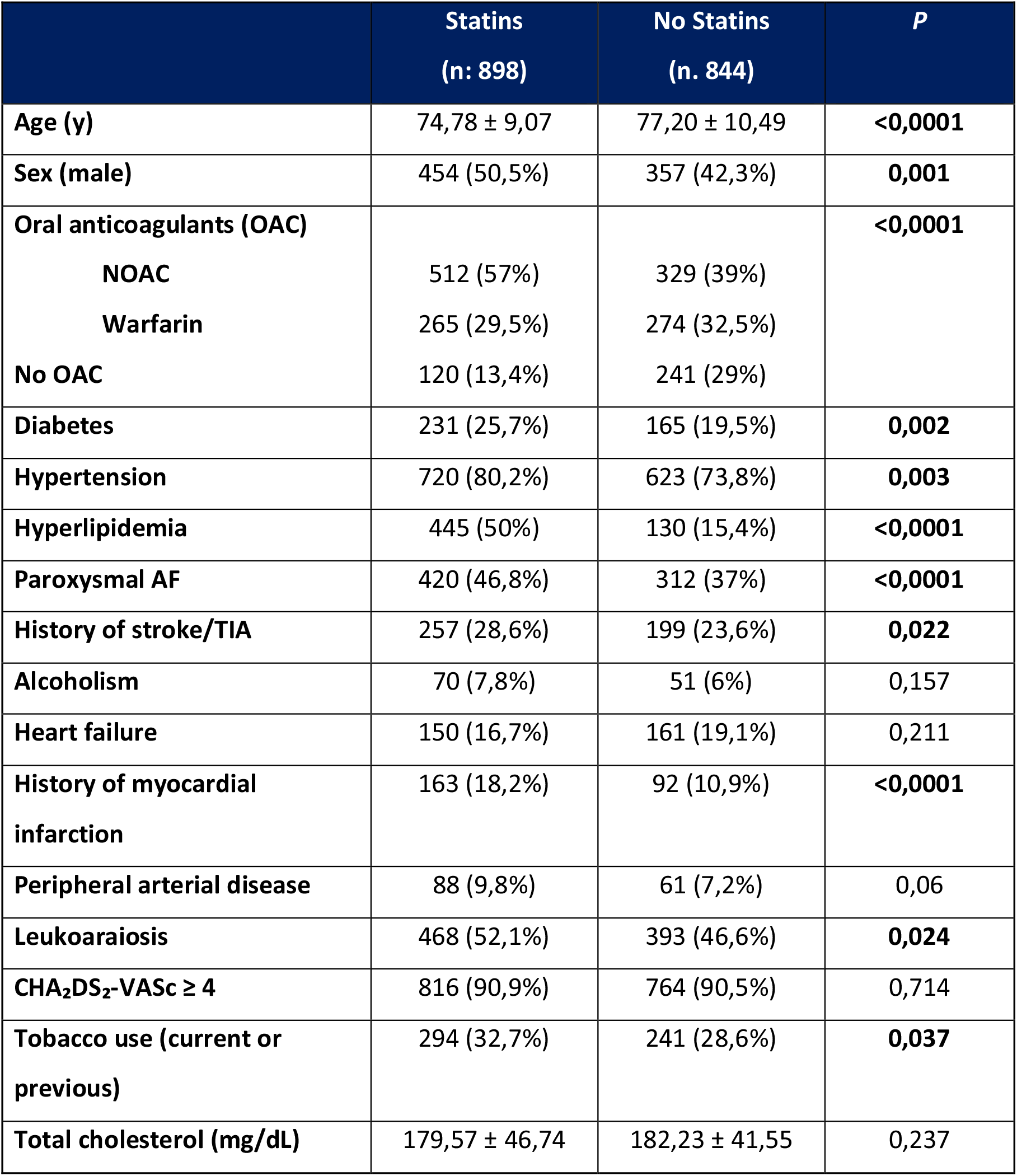
Univariate analysis: predictors for statin therapy.

**Table S2:** Univariate analysis: predictors for Combined Outcome.

|  | **Combined**<br>**Outcome**<br>**(n: 164)** | **No Combined**<br>**Outcome**<br>**(n. 1544)** | **P**<br>**(sign <0,05)** |
| --- | --- | --- | --- |
| **Age (y)** | 78,00 ± 8,92 | 75,71 ± 9,92 | **0,005** |
| **Sex (male)** | 72 (43,9%) | 720 (46,6%) | 0,511 |
| **Oral anticoagulants (OAC)** |  |  | **0,029** |
| **NOAC** | 50 (30,5%) | 759 (49,2%) |  |
| **Warfarin** | 56 (34,1%) | 483 (31,3%) |  |
| **No OAC** | 58 (35,4%) | 303 (19,6%) |  |
| **Diabetes** | 55 (33,5%) | 333 (21,6%) | **0,001** |
| **Hypertension** | 141 (86%) | 1178 (76,3%) | **0,006** |
| **Hyperlipidemia** | 54 (32,9%) | 512 (33,2%) | 1,000 |
| **Paroxysmal AF** | 58 (35,4%) | 662 (42,9%) | 0,067 |
| **History of stroke/TIA** | 44 (26,8%) | 403 (26,1%) | 0,852 |
| **Alcoholism** | 14 (8,5%) | 105 (6,8%) | 0,416 |
| **Heart failure** | 38 (23,2%) | 268 (17,4%) | 0,069 |
| **History of myocardial**<br>**infarction** | 30 (18,3%) | 218 (14,1%) | 0,162 |
| **Peripheral arterial disease** | 25 (15,2%) | 119 (7,7%) | **0,003** |
| **Leukoaraiosis** | 92 (56,1%) | 741 (48%) | 0,058 |
| **CHA<sub>2</sub>DS<sub>2</sub>-VASc ≥ 4** | 159 (97%) | 1389 (90%) | **0,002** |
| **Tobacco use (current or**<br>**previous)** | 48 (29,3%) | 479 (31%) | 0,721 |
| **Statin therapy** | 73 (44,5%) | 801 (51,9%) | 0,084 |

**Table S3:** Univariate Analysis on Statins and Combined and Secondary Outcomes.

|  | Statin Therapy<br>(n: 874) | No Statin Therapy<br>(n. 835) | *P* |
| --- | --- | --- | --- |
| **Combined Outcome** | 73 (8,4%) | 91 (10,9%) | 0,084 |
| **Ischemic Outcome** | 49 (5,6%) | 50 (6%) | 0,757 |
| **Hemorrhagic Outcome** | 25 (2,9%) | 45 (5,4%) | **0,010** |

## Notes

### Competing Interest Statement

The authors have declared no competing interest.

### Funding Statement

No external funding was received.

### Author Declarations

Ethical approval was obtained from local Institutional Review Boards as necessary.

